# Comparison of causal forest and regression-based approaches to evaluate treatment effect heterogeneity: An application for type 2 diabetes precision medicine

**DOI:** 10.1101/2022.11.07.22282023

**Authors:** Ashwini Venkatasubramaniam, Bilal A. Mateen, Beverley M Shields, Andrew T Hattersley, Angus G Jones, Sebastian J. Vollmer, John M. Dennis

## Abstract

**Objective:** To compare individualized treatment selection strategies based on predicted individual-level treatment effects from a causal forest machine learning algorithm and a penalized regression model.

**Study Design and Setting:** Cohort study characterizing individual-level glucose-lowering response (6 month reduction in HbA1c) in people with type 2 diabetes initiating SGLT2-inhibitor or DPP4-inhibitor therapy. Model development set comprised 1,428 participants in the CANTATA-D and CANTATA-D2 trials (SGLT2-inhibitor versus DPP4-inhibitor). For external validation, calibration of observed versus predicted differences in HbA1c in patient strata defined by size of predicted HbA1c benefit was evaluated in 18,741 UK primary care patients (Clinical Practice Research Datalink).

**Results:** Heterogeneity in treatment effects was detected in trial participants with both approaches (causal forest: 98.6% & penalized regression: 81.7% predicted to have a benefit on SGLT2-inhibitor therapy over DPP4-inhibitor therapy). In validation, calibration was good with penalized regression but sub-optimal with causal forest. A strata with an HbA1c benefit >10 mmol/mol with SGLT2-inhibitors (3.7% of patients, observed benefit 11.0 mmol/mol [95%CI 8.0-14.0]) was identified using penalized regression but not causal forest, and a much larger strata with an HbA1c benefit 5-10 mmol with SGLT2-inhibitors was identified with penalized regression (regression: 20.9% of patients, observed benefit 7.8 mmol/mol (95%CI 6.7-8.9); causal forest 11.6%, observed benefit 8.7 mmol/mol (95%CI 7.4-10.1).

**Conclusion:** When evaluating treatment effect heterogeneity researchers should not rely on causal forest (or other similar machine learning algorithms) alone, and must compare outputs with standard regression.

**What is new?:** *Question:* What is the comparative utility of machine learning compared to standard regression for identifying variation in patient-level outcomes (treatment effect heterogeneity) due to different treatments?

*Findings:* Causal forest and penalized regression models were developed using trial data to predict glycated hemoglobin [HbA1c]) outcomes with SGLT2-inhibitor and DPP4-inhibitor therapy in 1,428 individuals with type 2 diabetes. In external validation (18,741 patients), penalized regression outperformed causal forest at identifying population strata with a superior glycemic response to SGLT2-inhibitors compared to DPP4-inhibitors.

*Implications:* Studies estimating treatment effect heterogeneity should not solely rely on machine learning and should compare results with standard regression.

## Introduction

Randomized controlled trials (RCTs) are the gold standard for understanding the effect of treatments on clinical outcomes. Average treatment effects from RCTs are then used to support evidence-based clinical decision making for individual patients. This application of a population-level result to individual treatment selection may result in sub-optimal decision making, as the average treatment effects may only represent the individual experience of a subset of patients.^1^ As a result, there is great interest in developing precision medicine approaches to treatment, by characterizing patient sub-populations for which a treatment is most beneficial, or harmful. Such variability in patient level outcomes is known as treatment effect heterogeneity,^2,3^ and is often obscured by quoting average treatment effects. Importantly, if differences are clinically significant, characterizing treatment effect heterogeneity may allow specific treatments to be targeted at patients most likely to benefit.

Methods to evaluate treatment effect heterogeneity are not well established. One-variable-at-a-time subgroup analysis approaches have been shown to be rarely replicable due to low power, and will miss treatment effect heterogeneity induced by complex covariate relationships.^3^ Traditional regression-based models can be used to estimate treatment effect heterogeneity across multiple variables by defining potential treatment-covariate interactions for each covariate of interest, but require these covariates to be specified by the analyst. Results may in particular be subject to the risk of Type I Error rate inflation (false positives) with small sample sizes, which may not be solved by penalized or shrinkage methods.^4^ Recently, machine learning algorithms, in particular causal forest, have been developed to specifically assess treatment effect heterogeneity and represent a data-driven alternative to regression-based approaches.^5,6^ Whilst such machine learning approaches have been demonstrated to overcome challenges associated with reliance on manual input, their comparative utility relative to regression-based approaches for the purposes of treatment selection based on treatment effect heterogeneity has not previously been assessed.^7^

This issue of sub-optimal (personalized) decision making is potentially evident in the pharmacological management of Type 2 diabetes; a heterogenous chronic condition with multiple treatment options prescribed with the primary clinical purpose of lowering blood glucose (glycated hemoglobin [HbA1c]) levels. SGLT2-inhibitors (SGLT2-i) and DPP4-inhibitors (DPP4-i) are two commonly prescribed glucose-lowering treatment options,^8^ recommended after metformin in type 2 diabetes clinical guidelines.^9^ Whilst RCT data suggest that the glucose-lowering efficacy of both treatments is on average similar,^10^treatment effect heterogeneity is plausible due to the marked variation in the clinical characteristics of people with type 2 diabetes, and the two drugs’ differing mechanisms of action.^11^ As such, our primary objective in this study was to compare individualized treatment selection strategies based on predicted treatment effects from a causal forest algorithm and a penalized regression model, using the clinically relevant context of selecting between SGLT2-i and DPP4-i therapy for people with type 2 diabetes.

## Methods

### Overview

Two treatment effect heterogeneity models (causal forest and penalized regression) were developed to predict HbA1c-lowering efficacy with SGLT2-i and DPP4-i therapy using individual-level participant data from two large RCTs. Performance of individualized treatment selection strategies derived from each model was evaluated in routine clinical data.

### Data sources & Handling

#### Clinical trial data (development dataset)

Individual participant data from 2 active comparator glucose-lowering efficacy RCTs of SGLT2-i (Canagliflozin) and DPP4-i (Sitagliptin) therapy (2010-2012) in people with type 2 diabetes were accessed from the Yale University Open Data Access Project (https://yoda.yale.edu/). Data on participants randomized to either SGLT2-i or DPP4-i in the CANTATA-D and CANTATA-D2 were pooled for analysis; these trials differed only in background glucose-lowering therapy not in any other inclusion criteria. Trial results to compare the average HbA1c-lowering efficacy of the two therapies have been previously published.^12,13^

#### Routine clinical data (test dataset)

Anonymized primary care electronic health records were extracted from UK Clinical Practice Research Datalink (CPRD) GOLD.^14^ New users of SGLT2-i and DPP4-i therapies (i.e. patients initiating one of these therapies for the first time) after January 1st, 2013, were identified, following our previously published protocol.^15^ We then excluded patients prescribed a SGLT2-i or DPP4-i as first-line treatment (as this is not in-line with treatment guidelines),^9^ patients co-treated with insulin, patients with eGFR <45 (where SGLT2-i prescription is usually contraindicated), patients with a missing baseline HbA1c or a baseline HbA1c <53 or _≥_ 120 mmol/mol (with baseline defined as the closest HbA1c to drug initiation within –91/+7 days).

#### Predictors

Across both sources, the following clinical features were extracted for each individual: initial HbA1c, age at treatment, sex, estimated glomerular filtration rate (eGFR), Alanine Aminotransferase (ALT), body mass index (BMI), High-density lipoprotein cholesterol (HDL-c), High-density lipoprotein cholesterol (HDL-c), Triglycerides, Albumin, and Bilirubin. These features were selected due to their availability in a majority of individuals in both the trial and routine data.

Diabetes duration was redacted from the RCT data so was not evaluated. In CPRD, where a systematic baseline assessment was not available, we used the most recent value in the 2 years prior to drug initiation available in the primary care record. In CPRD, we also identified the number of currently prescribed glucose-lowering treatments, and the number of glucose-lowering drug classes ever prescribed, as addition patient-level confounding factors.

#### Missing Data Handling

In the trials, missing values in all the covariates were imputed using missForest, a random forest based imputation method.^16^ For validation of the model developed in the trials in CPRD, we conducted complete case analysis, as missing values were considered likely to be missing not at random.^17^

#### Statistical Modelling

Two treatment effect heterogeneity models were developed using RCT (training) data. During model development the prediction target was the achieved HbA1c 6 months after drug initiation (a measure of glucose-lowering efficacy), presented as a continuous measure. In the trials, this was defined as the last-observation-carried-forward HbA1c from 3-months if the 6-month value was not available. In CPRD, this was defined as the closest HbA1c to 6 months (within 3-15 months) after initiation, on unchanged glucose-lowering therapy. Subsequently, utility of the models for selecting optimal treatment for patients was evaluated in routine clinical electronic medical record data using a novel framework.^11^.

#### Model development in trial data: Penalized regression

A multivariable linear regression model was fitted to the training dataset composed of allbaseline features (see ***Predictors***), the outcome and the treatment indicator. Each of the eleven continuous baseline features was modelled as a 3-knot restricted cubic spline to allow for non-linearity. Interaction term for each baseline feature:treatment indicator pair were included to estimate treatment effect heterogeneity. No variable selection was applied, but optimal penalty factors, based on AIC, were estimated separately for main effects, non-linear effects, and interaction terms, using a ridge regression approach (*pentrace* function in R package *RMS*).^18^ Optimism-adjusted model fit (R^2^), root mean square error (RMSE), and the calibration slope and calibration-in-the-large were estimated, although these test the ability of a model to predict the outcome, and are therefore of limited use when evaluating treatment effect heterogeneity. Relative feature importance, in terms of treatment effect heterogeneity, was assessed by ranking features by the proportion of chi-squared explained by the interaction term for that feature, with bootstrapped confidence intervals.

#### Model development in trial data: Causal forest

A causal forest model was also fitted over the training dataset. The causal forest model was built over 5000 causal trees and used default tuning parameters for growing the many tree structures. Tuning parameters used for growing an individual causal tree included setting a minimum of ten patients within a determined subgroup and splitting the training dataset equally into two separate samples for first determining the tree structure, and then utilising the second sample for treatment effect estimation at each determined subgroup. Variable importance measures computed from trees in the forest highlight the covariates selected most frequently by the model. However, CART and associated ensemble structures (e.g., random forests) have been shown to be biased towards splitting over covariates that offer many potential values to split on (e.g., continuous covariates) as compared to covariates with few categories (e.g., binary covariates). To account for this problem of biased variable selection, adjusted feature importance in the form of p-values were determined using a permutation-based test.^19,20^ A p-value for each covariate is computed by determining the proportion for which importance measures from forest models over permuted responses are greater than the measure obtained for a forest using an unpermuted response.

#### Model evaluation in routine clinical data

Utility of the two treatment effect heterogeneity modelling approaches for selecting the likely most effective therapy for patients was tested in CPRD. The first step was to estimate the difference in the in predicted HbA1c outcome (the conditional average treatment effect; see Box 1) for each patient using both models. The accuracy of the CATE cannot be evaluated at the patient-level (as patients receive either SGLT2-i or DPP4-i but not the other). However, it can be used to define and test a treatment selection decision rule in patient strata defined by the difference in predicted HbA1c outcome, as follows: For each model, the difference in HbA1c outcome was estimated for each patient. For penalized regression this was the difference in predicted HbA1c outcome on the two therapies. In the causal forest algorithm, the difference in HbA1c outcome is explicitly estimated. Strata were then defined by defined by decile of predicted difference in predicted HbA1c outcome, and by clinically defined HbA1c cut-offs of predicted difference in HbA1c outcome (SGLT2i benefit: _≥_10, 5-10, 3-5, 0-3 mmol/mol; DPP4i benefit: _≥_5, 3-5, 0-3 mmol/mol). To compare performance of each model, we tested whether within-strata HbA1c outcome differences were consistent with predictions. Linear regression models were used to contrast HbA1c outcome in concordant (i.e. therapy received is the therapy predicted to have greatest HbA1c lowering) versus discordant (i.e. therapy received is the predicted non-optimal therapy) subgroups. As CPRD patients were not randomized to treatment, models were adjusted for all features used in the treatment selection model, and confounding factors (see *Predictors*). Statistical analysis used R software, with causal forest fitted using the *grf* package.^21^

##### Box 1: Primer on Conditional Average Treatment Effect (CATE) Estimation

###### Evaluation framework

In a potential outcomes framework, the causal effect of a treatment on a patient is defined by the difference in outcomes, where the outcomes are obtained for two different treatment assignments. The conditional average treatment effect (CATE) is defined as the average over individual treatment effects for a subpopulation determined by specific patient characteristics. The estimation of such subgroup-specific treatment effects has traditionally relied on a manual comparison of pre-defined patient sub-populations. However, this is not necessarily possible for subgroups determined by unknown covariate relationships or for higher-dimensional datasets. We evaluate two different methods that are able to estimate conditional average treatment effects, which represent differential patient responses to a treatment allocation.

###### Penalized regression

Standard maximum likelihood regression models can estimate CATE by including treatment-by-covariate interaction terms. For each covariate, the interaction term coefficient(s) represent the estimated differential treatment effect associated with that covariate. The model can then be used to predict the counterfactual outcome on each therapy, conditional on the features included as interaction terms. The difference between the predicted outcome on each therapy provides an estimate of the patient-level treatment effect. Penalized regression can be used to reduce overfitting and potentially improve prediction in new data.

###### Causal forest

Causal forest is a data-driven ensemble method built over many individual causal trees to estimate the CATE.^6^ A causal tree^5^ modifies the traditional CART structure^22^ to explicitly optimise for treatment effect heterogeneity and generates estimates at the leaves of the trees. Causal trees utilise a separate sample to detect the tree structure and another sample to estimate the treatment effects, this double-sample approach (also referred to as honest) helps to overcome the problem of over-fitting. Similar to the random forest for outcome prediction, each causal tree within the causal forest is built over a bootstrap sample from the training data and the forest averages over the tree generated treatment effects. In general, a forest over a large number of individual trees has been shown to more stable and produce more accurate results than an individual tree.^19^

## Results

### Participant cohort

Baseline clinical characteristics of the trial cohort used for model development (n=1,428) are reported in **Table 1**. 61 participants were excluded as they had no on-treatment HbA1c outcome available (**sFlowchart 1**). Mean achieved HbA1c at 26 weeks was 53.0 (SD 9.8) on SGLT2-i and 54.1 (SD 10.9) on DPP4-i.

**Table 1:**
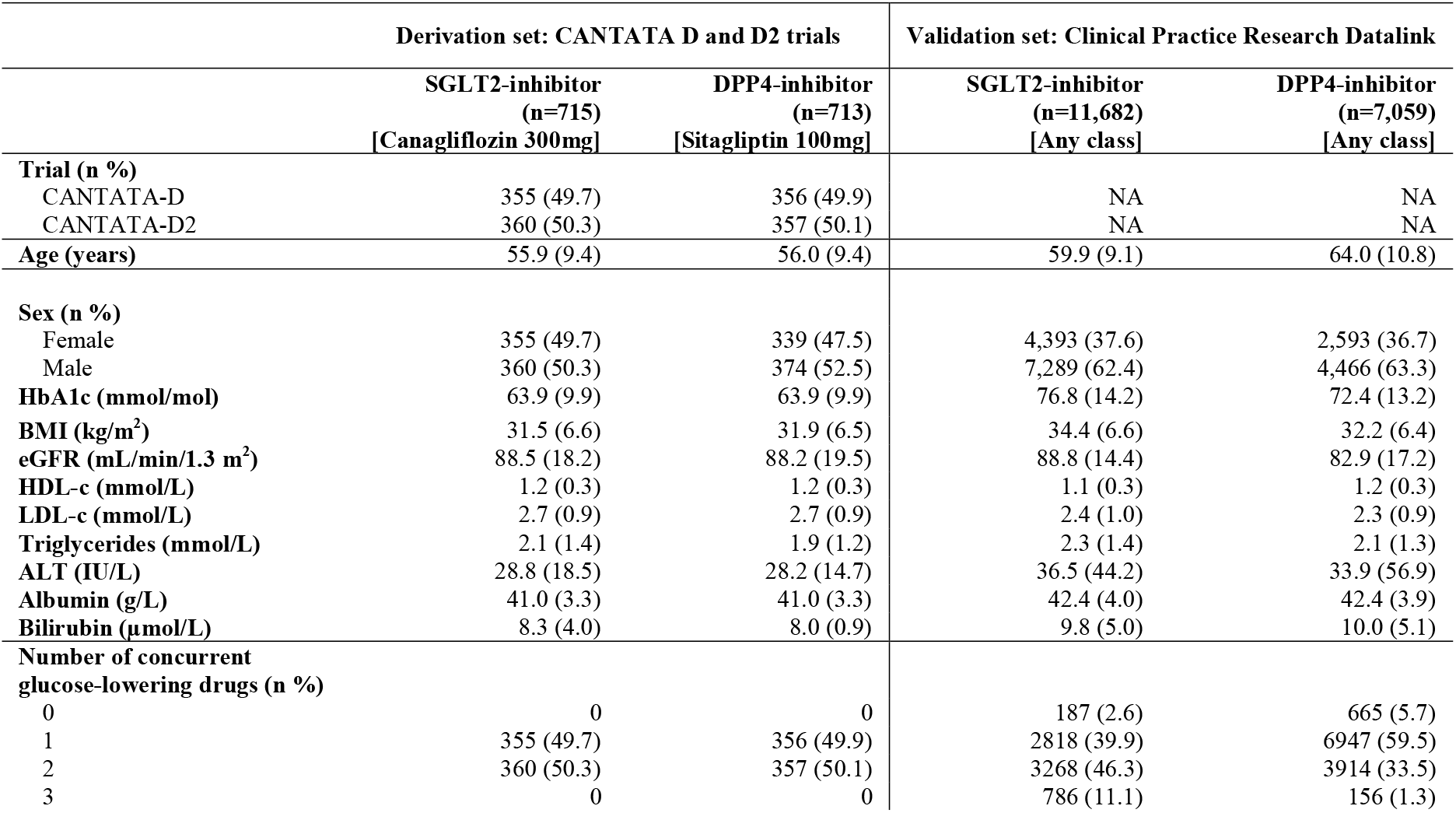
Baseline clinical characteristics by initiated drug class in CANTATA D and D2 trials, and CPRD. Data are mean (SD) unless stated

### Model development

#### Penalized regression

In the development cohort the median average treatment effect was estimated as a 1.9 (IQR 0.5, 3.6) greater HbA1c reduction with SGLT2-i compared to DPP4-I (**sFigure 1a**). There was evidence of heterogeneity of treatment effect with a predicted greater HbA1c reduction with SGLT2-i versus DPP4-i for 1,216 (81.7%) of trial participants. Optimism-adjusted model performance statistics for predicting HbA1c outcome were: RMSE 8.1 (95%CI 7.6, 8.1) mmol/mol, R^2^ 0.30 (95%CI 0.26, 0.36), calibration slope 0.98 (95%CI 0.98, 1.00), calibration in the large 0.86 (−0.19. 0.95).

#### Causal forest

The median average treatment effect in the development cohort was a 1.6 (IQR 0.6, 2.5) greater HbA1c reduction with SGLT2-i therapy (**sFigure 1b**). There was evidence of heterogeneity in individual treatment effects (p=0.005), although 1,408 (98.6%) of participants were predicted to have a greater benefit on SGLT2-i therapy.

#### Model specification

#### Most influential predictors of differential treatment effect

Figure 1. reports the most influential predictors for differential treatment effect for the regression and causal forest approaches. Baseline HbA1c, age, ALT and triglycerides were the top 4 predictors identified by both approaches.

**Figure 1:**
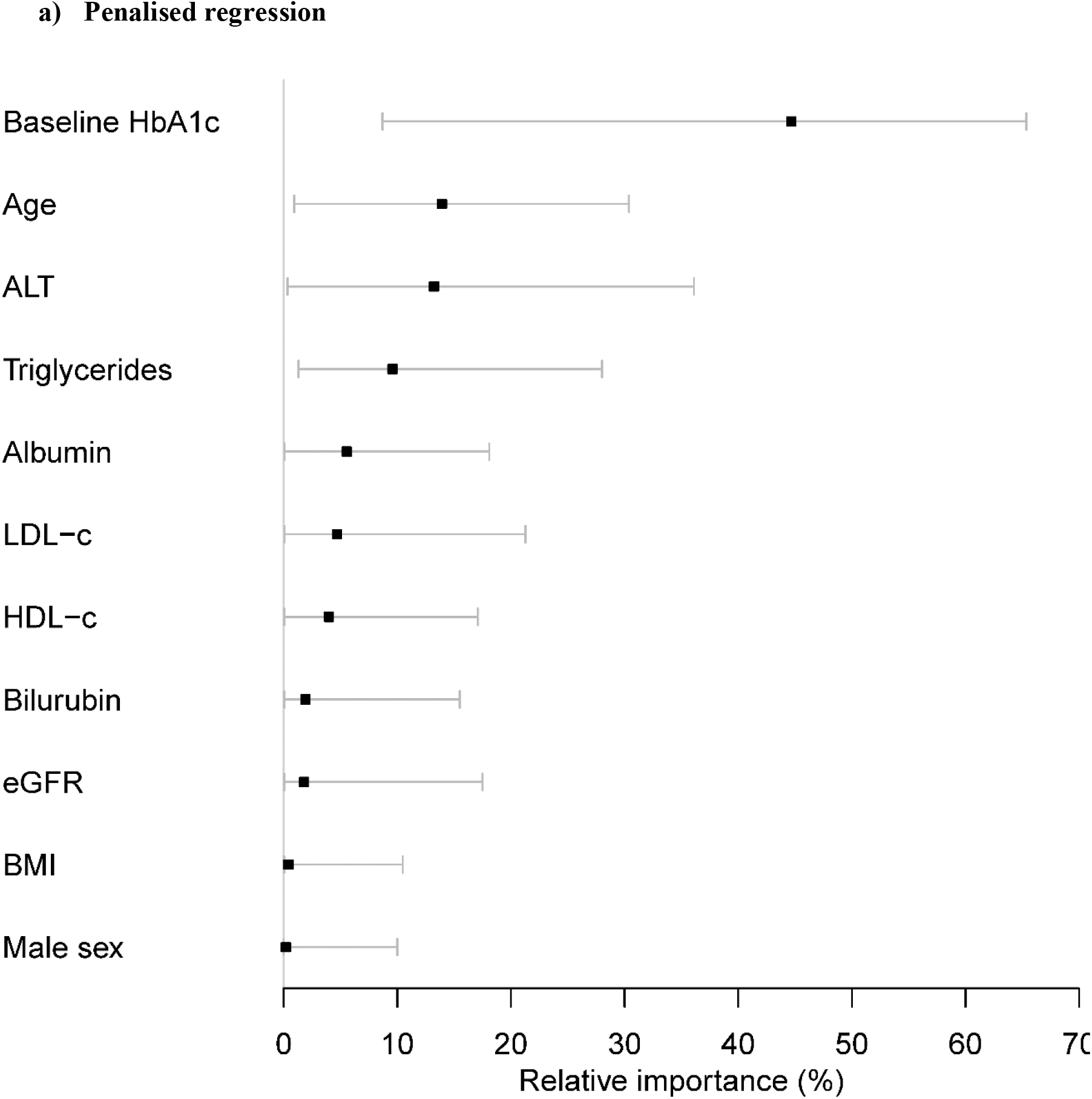
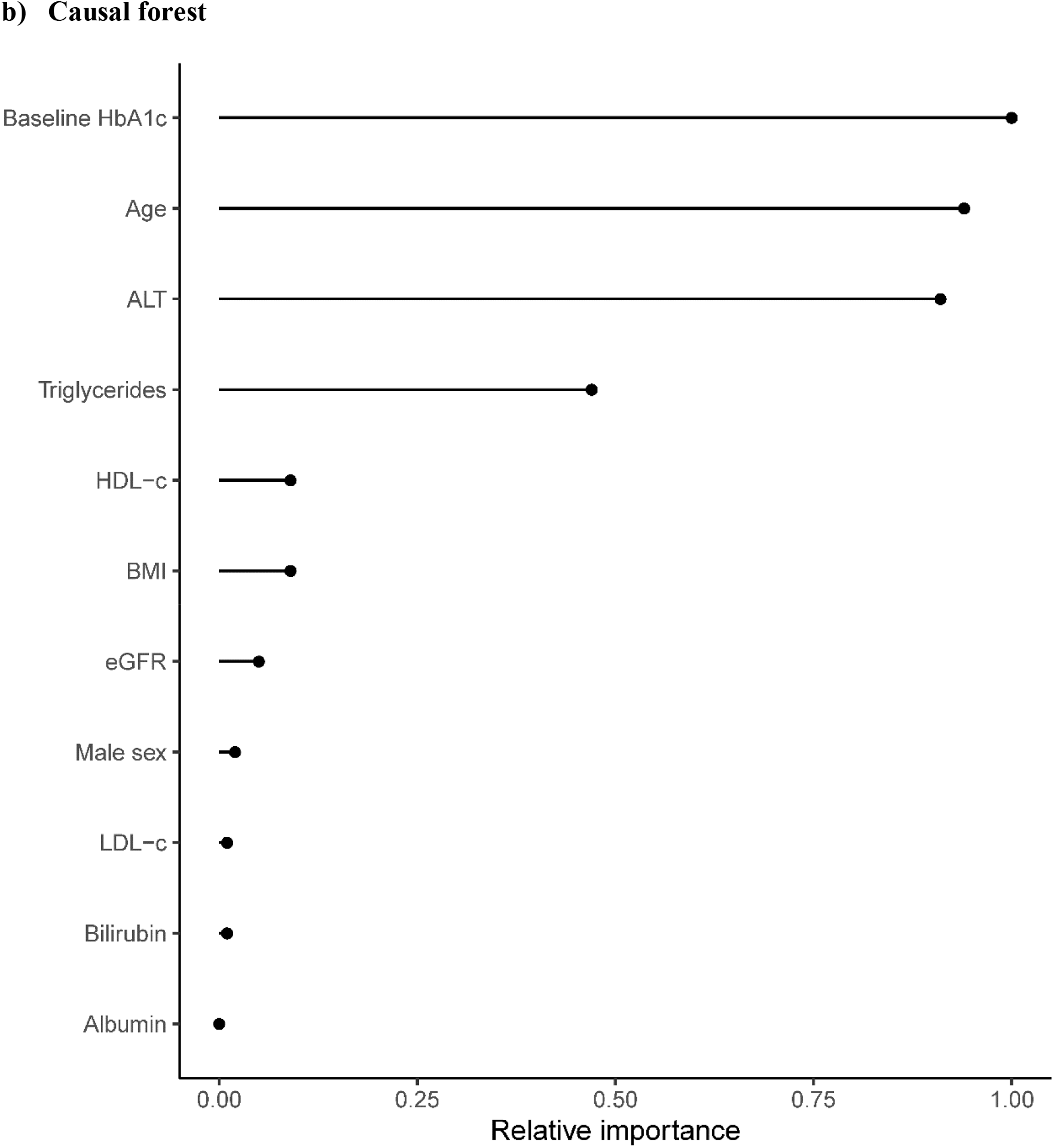
Relative feature importance for treatment selection between SGLT2-inhibitor and DPP4-inhibitor treatment, for all clinical features. a) Penalized regression. Feature importance reflects the proportion of chi-squared explained by drug-by-covariate interaction terms for each clinical feature in multivariable analysis, as these represent differential treatment effects for the two therapies. Bars represent bootstrapped 95% confidence intervals. **b) Causal forest model**. Adjusted importance (using p-values) represent the covariates selected most often by trees within the causal forest, after controlling for biased variable selection. Permutation-based tests generate p-values for each covariate, using an understanding that spurious splits in trees would continue to occur in the presence of a permuted outcome unless these splits also reflect the true underlying association. For the purpose of comparison, inverse p-values are presented as relative importance measures.

#### Model external validation: performance for treatment selection in routine clinical data

Utility for selecting treatment was evaluated in 18,741 patients initiating DPP4-i (n=11,682), or SGLT2-i (n=7,059) in CPRD (**sFlowchart**). Patients initiating each therapy differed in all clinical characteristics except sex and baseline albumin (**Table 1**). In particular, patients initiating DPP4-i were on average older than those initiating SGLT2-i (mean 64.0 versus 59.9 years), had a lower baseline HbA1c (mean 72.4 versus 76.8 mmol/mol), and had lower BMI (mean 32.2 versus 24.4 kg/m^2^) and eGFR (mean 82.9 versus 88.8 mL/min/1.3 m^2^

The distribution of model predicted treatment difference for the regression and causal forest approaches are shown in **Figure 2**. The regression model predicted that 87% (n=16,276) of patients would benefit on SGLT2-i and 13% (n=2,465) on DPP4-i. In contrast, the causal forest model predicted that nearly all patients (99.7% [n=18,689]) would benefit on a SGLT2-i.

**Figure 2:**
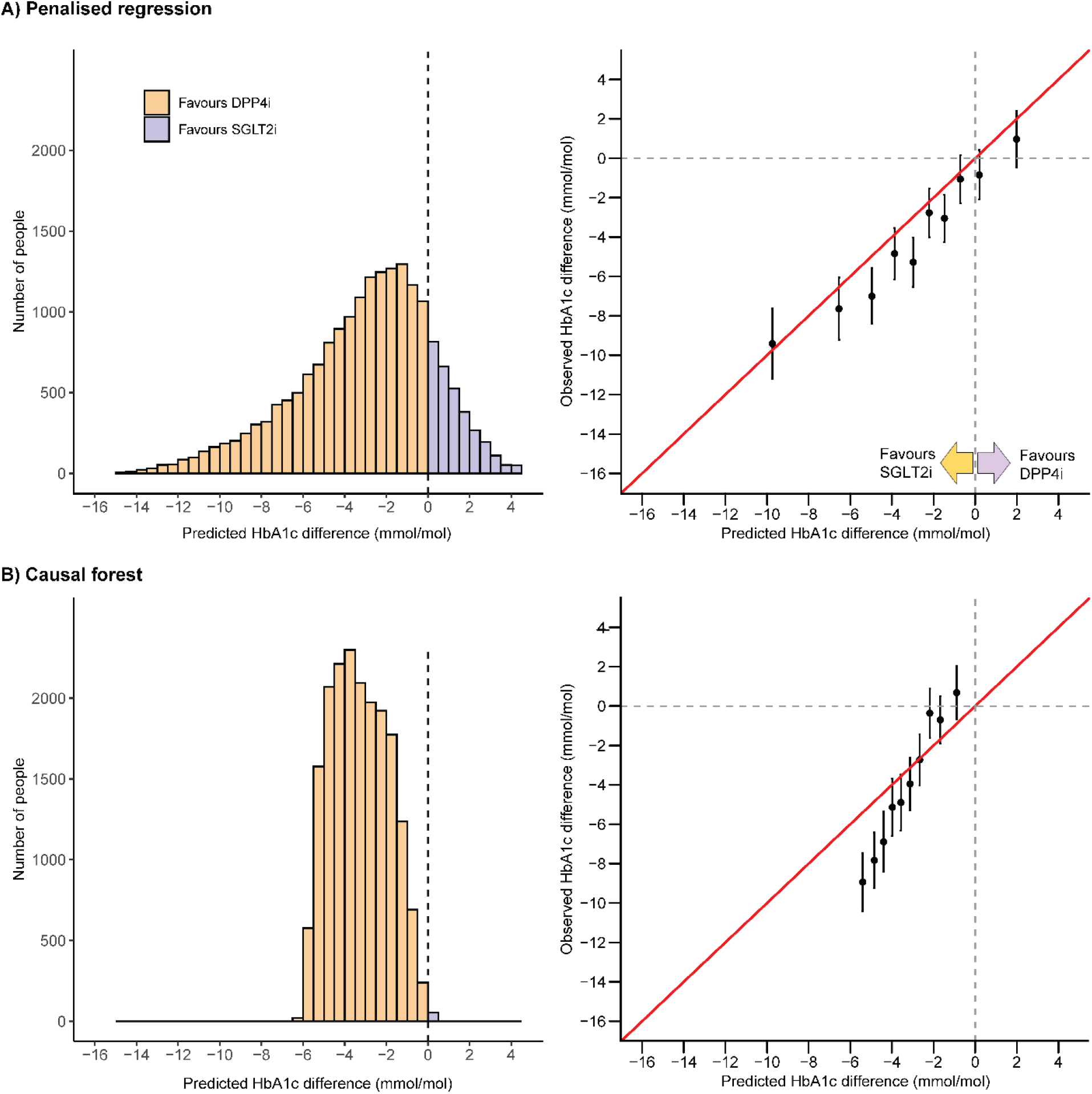
Final treatment selection model performance for A) Penalized regression and B) Causal forest in CPRD validation data. Left panels show the distribution of predicted individualized treatment effects. Negative values reflect a predicted benefit on SGLT2-inhibitor treatment, positive values reflect a predicted HbA1c benefit on DPP4-inhibitor treatment. Right panels show calibration between observed and predicted treatment effects, across strata defined by decile of predicted treatment effect. Estimates are adjusted for clinical features in the treatment selection model to improve precision and control for potential differences in covariate balance within strata.

From the regression model there was good calibration between observed and predicted estimates, across deciles of predicted treatment effect (**Figure 2**). This included reliably identifying the smaller group of patients with a predicted treatment benefit on DPP4-i. Although the causal forest model did reliably identify patients with differences in observed treatment effect, the model did not show good calibration (**Figure 2**). The causal forest predicted treatment effects were in a much narrower range than observed treatment effects, and the model did not identify a patient strata with an observed treatment benefit on DPP4-i. In strata defined by clinical cut-offs for predicted treatment benefit (**Table 2**), the regression model reliably identified 687 (3.7%) patients with a marked (_≥_10 mmol/mol) observed benefit on SGLT2-i. This group was not identified using the causal forest model. The regression model also identified a much larger group of patients with an observed benefit with SGLT2-i of 5-10 mmol/mol (n=3,920 [20.9%]) compared to the causal forest model (n=2,175 [11.6%]). Similarly, a group with a >3mmol/mol benefit on DPP4-i was identified with the regression model (n=270 [1.4%]) but not the causal forest.

**Table 2:** External validation in CPRD: Observed treatment effects across strata defined by clinical cut-offs of predicted treatment benefit. Estimates are adjusted for clinical features in the treatment selection model (to improve precision and control for potential differences in covariate balance within subgroups).

**Penalized regression model external validation**
| Predicted HbA1c difference | Observed treatment difference (mmol/mol; negative favors SGLT2-i) |  |  |  |  |
| --- | --- | --- | --- | --- | --- |
|  | N patients | Treatment difference | Lower CI | Upper CI | p-value |
| **Overall** | 18,741 | -4.5 | -4.9 | -4.0 | <0.001 |
| **Strata** |  |  |  |  |  |
| SGLT2-i benefit by any mmol/mol | 15626 | -5.1 | -5.5 | -4.6 | <0.001 |
| SGLT2-i benefit by $\geq 10$ mmol/mol | 687 | -11.0 | -14.0 | -8.0 | <0.001 |
| SGLT2-i benefit by 5-10 mmol/mol | 3920 | -7.8 | -8.9 | -6.7 | <0.001 |
| SGLT2-i benefit by 3-5 mmol/mol | 3763 | -5.4 | -6.3 | -4.4 | <0.001 |
| SGLT2-i benefit by 0-3 mmol/mol | 7256 | -2.6 | -3.2 | -2.0 | <0.001 |
| DPP4-i benefit by any mmol/mol | 3115 | 0.2 | -0.8 | 1.2 | 0.700 |
| DPP4-i benefit by 0-3 mmol/mol | 2845 | 0.0 | -1.1 | 1.1 | 0.983 |
| DPP4-i benefit by $\geq 3$ mmol/mol | 270 | 3.1 | -1.5 | 7.7 | 0.186 |

**Causal forest external validation**
| Predicted HbA1c difference | Observed treatment difference (mmol/mol; negative favors SGLT2-i) |  |  |  |  |
| --- | --- | --- | --- | --- | --- |
|  | N patients | Treatment difference | Lower CI | Upper CI | p-value |
| **Overall** | 18741 | -4.5 | -4.9 | -4.0 | 0.003 |
| **Strata** |  |  |  |  |  |
| SGLT2-i benefit by any mmol/mol | 18689 | -4.5 | -4.9 | -4.0 | <0.001 |
| SGLT2-i benefit by $\geq 10$ mmol/mol | 0 | NA | NA | NA | NA |
| SGLT2-i benefit by 5-10 mmol/mol | 2175 | -8.7 | -10.1 | -7.4 | <0.001 |
| SGLT2-i benefit by 3-5 mmol/mol | 8676 | -5.8 | -6.5 | -5.2 | <0.001 |
| SGLT2-i benefit by 0-3 mmol/mol | 7838 | -1.0 | -1.6 | -0.4 | 0.001 |
| DPP4-i benefit by any mmol/mol | 52 | 2.0 | -12.7 | 16.8 | 0.78 |
| DPP4-i benefit by 0-3 mmol/mol | 52 | 2.0 | -12.7 | 16.8 | 0.78 |
| DPP4-i benefit by $\geq 3$ mmol/mol | 0 | NA | NA | NA | NA |

## Discussion

Our study provides a comparison of causal forest and regression approaches to detect and characterize treatment effect heterogeneity, as well as to operationalize it for treatment selection. Specifically, we observed that while both approaches detect treatment effect heterogeneity in glucose-lowering efficacy for SGLT2-i and DPP4-i, this translates into marked differences in predicted treatment benefit for individual patients. Through external validation using real-world (routinely collected) data, we establish the utility of both approaches for identifying strata with an observed benefit on one treatment over the other. We found a regression-based model performed substantially better than causal forest for identifying strata with a clinically important observed treatment benefit on SGLT2-i compared to DPP4-i. In contrast to causal forest, the regression model was also able to identify a smaller strata with a likely observed treatment benefit on DPP4-i.

From a methodological perspective, the analysis adds to the growing literature showing limited, if any, performance improvement for machine learning over regression in tasks utilizing structured clinical data,^23-26^ although our study provides important new evidence as previous evaluations have focused on performance for risk prediction rather than treatment effect heterogeneity. Interestingly, in this setting we found the causal forest algorithm outputted substantially more conservative estimates of treatment effect heterogeneity compared to penalized regression. Although we demonstrate this with only a single outcome in a limited trial population, this reflects precisely the type of clinical dataset where such data-driven methods for treatment effect heterogeneity are increasingly being deployed, for example in evaluation of risk of harm of intensive blood pressure management in the SPRINT trial,^27^ and evaluation of heterogeneity in mortality risk in people with diabetes in the ACCORD trial.^28^ Given the lower performance of the causal forest algorithm in external validation, our study suggests that further research is urgently needed to understand the reasons underlying differences in outputs from treatment effect heterogeneity focused machine learning and regression based approaches in relatively low dimensional health datasets. In the meantime, we recommend that, when evaluating treatment effect heterogeneity, researchers do not rely on causal forest (or other similar machine learning) algorithms alone and compare outputs with standard regression. This is further supported by recent work suggesting subgroups defined by heterogenous treatment effects using causal trees may not be reproducible across randomized trials.^29^

Moreover, in the specific context of type 2 diabetes management, our results support recent work showing that a ‘precision’ approach to treatment is possible by demonstrating clinically relevant heterogeneity of treatment response that can be predicted using simple patient characteristics and routine biomarker tests.^12,13^ Our findings raise the possibility of targeting specific treatment, to patients most likely to have a greater HbA1c response, using characteristics that are already routinely measured. However, a limitation is that we evaluated only a single outcome, HbA1c. Treatment decisions are multi-factorial, and potential glycemic benefit should be considered alongside differences in side-effect profile, likely tolerability, and cardiovascular and renal benefit, and a similar approach to stratifying risk of these outcomes based on patient characteristics may be feasible in future.^11,30^

Strengths of our study include the systematic comparison of both modelling approaches in the same datasets, and the use of individual-level trial data to develop treatment effect heterogeneity models, meaning randomization may allow a causal interpretation of individual-level treatment effects.^31^ Whilst research to develop optimal methods for predicting treatment effect heterogeneity, and to evaluate their performance, has been called for in the recent PATH statement,^2^ the evaluative framework applied in this study can be applied for any future study aiming to evaluate the value of using patient level features to inform a precision medicine approach to treatment in any disease with multiple treatment options.^11^

A limitation of our study is that we only compared performance in a single, low dimensional setting with a continuous outcome; it is conceivable that causal forest may outperform regression-based approaches with high dimensional or less structured data than those captured in clinical trial and routine clinical data. A further limitation is that we only evaluated a single machine learning approach. Causal forest was chosen as it is widely used with easy to use software available. We cannot comment on the performance of other treatment effect heterogeneity focused algorithms, such as the LASSO,^32^ Bayesian frameworks,^33-35^ and a generic machine learning approach, that were not evaluated. Finally, as our validation dataset was observational, we cannot rule out unmeasured confounding as a potential explanation for our findings.^36^

## Conclusions

The causal forest machine learning algorithm is outperformed by standard regression when identifying patients with a treatment benefit of one blood glucose-lowering drug over another. Given the rapidly growing interest in precision medicine, further research is urgently needed to understand the settings in which different classical and data-driven modelling approaches can be effectively deployed to reliably detect and quantify treatment effect heterogeneity.

## Data Availability

No additional data are available from the authors although CPRD data are available by application to CPRD Independent Scientific Advisory Committee, and the clinical trial data are accessible via application from the Yale University Open Data Access Project.

## Acknowledgements

This article is based in part on data from the Clinical Practice Research Datalink obtained under license from the UK Medicines and Healthcare products Regulatory Agency. CPRD data is provided by patients and collected by the NHS as part of their care and support. This study, carried out under YODA Project # 2017-1816, used data obtained from the Yale University Open Data Access Project, which has an agreement with JANSSEN RESEARCH & DEVELOPMENT, L.L.C.. The interpretation and reporting of research using this data are solely the responsibility of the authors and does not necessarily represent the official views of the Yale University Open Data Access Project or JANSSEN RESEARCH & DEVELOPMENT, L.L.C..

## Ethics/data approvals

Approval and data access for the study was granted by the CPRD Independent Scientific Advisory Committee (ISAC 13_177R), and the YODA Project (# 2017-1816).

## Authors’ contributions

JMD, SJV, BAM, and AV designed the study. JMD and AV undertook the analysis in the RCT data, JMD ran the analysis in CPRD. AV and JMD drafted the article. All authors provided support for the analysis and interpretation of results, critically revised the article, and approved the final article.

## Competing interests

BAM is an employee of the Wellcome Trust and holds an honorary post at University College London for the purposes of carrying out independent research; the views expressed in this manuscript do not necessarily reflect the views of the Wellcome Trust. SJV declares funding from IQVIA. All other authors declare no competing interests.

### Prior Presentation

None

