## Supplementary material for "Comparison of causal forest and regression-based approaches to evaluate treatment effect heterogeneity: An application for type 2 diabetes precision medicine": Data supplement

### sFlowchart

#### A) CANTATA D and D2 trials (development cohort)

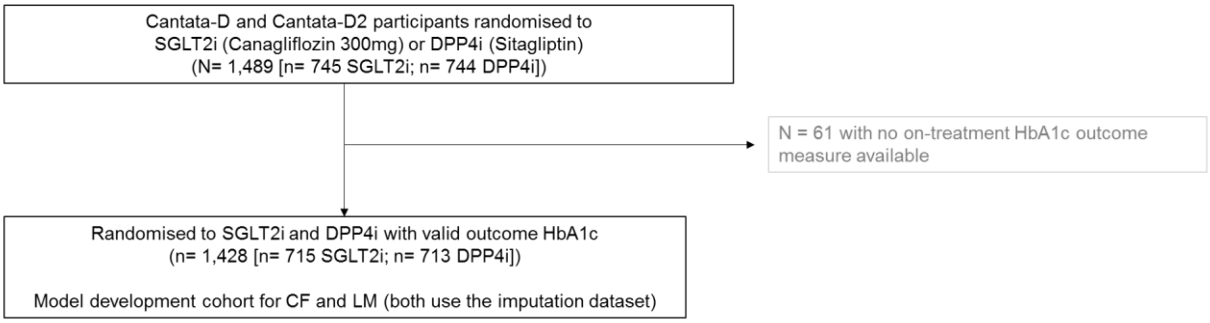


#### B) CPRD patient flow and inclusion criteria (validation cohort)

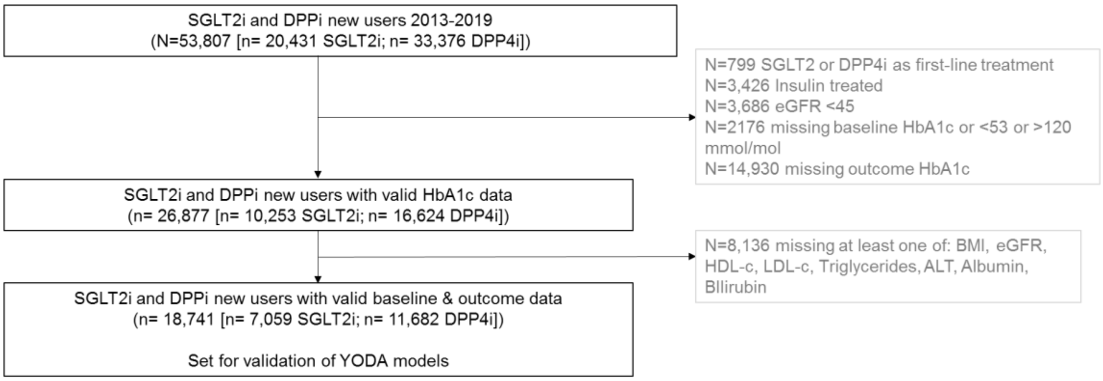

**sFigure 1: Distribution of the predicted individualized treatment effect of SGLT2-inhibitor treatment compared to DPP4-inhibitor treatment in the RCT derivation data. a) Penalized regression.** SGLT2-i was the predicted optimal therapy for 1,216 (85.1%) participants, DPP4-i for 212 (14.8%) participants. **b) Causal forest.** SGLT2-i was the predicted optimal therapy for 1,414 (99.0%) participants, DPP4-i for 14 (1.0%) of participants

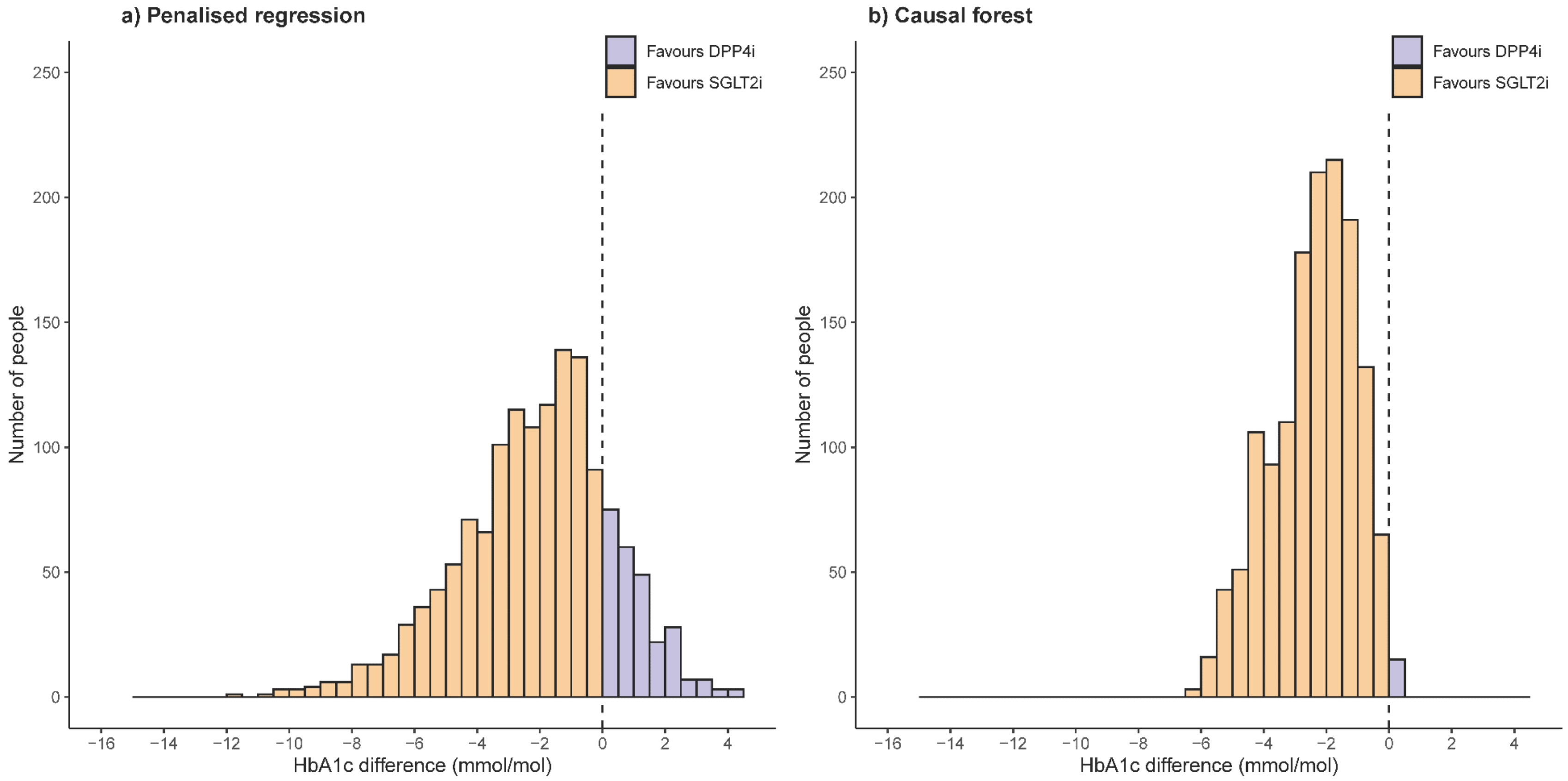
